# AI-Assisted Data Extraction with a Large Language Model: A Study Within Reviews

**DOI:** 10.1101/2025.03.20.25324350

**Authors:** Gerald Gartlehner, Shannon Kugley, Karen Crotty, Meera Viswanathan, Andreea Dobrescu, Barbara Nussbaumer-Streit, Graham Booth, Jonathan R. Treadwell, Jung Min Han, Jesse Wagner, Eric A. Apaydin, Erin L. Coppola, Margaret Maglione, Rainer Hilscher, Robert Chew, Meagan Pilar, Bryan Swanton, Leila C. Kahwati

## Abstract

**Background:** Data extraction is a critical but error-prone and labor-intensive task in evidence synthesis. Unlike other artificial intelligence (AI) technologies, large language models (LLMs) do not require labeled training data for data extraction.

**Objective:** To compare an AI-assisted to a traditional y data extraction process.

**Design:** Study within reviews (SWAR) utilizing a prospective, parallel group comparison with blinded data adjudicators.

**Setting:** Workflow validation within six ongoing systematic reviews of interventions under real-world conditions.

**Intervention:** Initial data extraction using an LLM (Claude versions 2.1, 3.0 Opus, and 3.5 Sonnet) verified by a human reviewer.

**Measurements:** Concordance, time on task, accuracy, recall, precision, and error analysis.

**Results:** The six systematic reviews of the SWAR contributed 9,341 data elements, extracted from 63 studies. Concordance between the two methods was 77.2%. The accuracy of the AI-assisted approach compared with enhanced human data extraction was 91.0%, with a recall of 89.4% and a precision of 98.9%. The AI-assisted approach had fewer incorrect extractions (9.0% vs. 11.0%) and similar risks of major errors (2.5% vs. 2.7%) compared to the traditional human-only method, with a median time saving of 41 minutes per study. Missed data items were the most frequent errors in both approaches.

**Limitations:** Assessing the concordance of data extractions and classifying errors required subjective judgment. Tracking time on task consistently was challenging.

**Conclusion:** The use of an LLM can improve accuracy of data extraction and save time in evidence synthesis. Results reinforce previous findings that human-only data extraction is prone to errors.

**Primary Funding Source:** US Agency for Healthcare Research and Quality, RTI International

**Registration:** SWAR28 Gerald Gartlehner (2023 FEB 11 2102).pdf

## Introduction

Among all steps involved in conducting systematic reviews, data extraction (i.e., the process of extracting data from primary studies into standardized tables) is one of the most critical, labor-intensive, and error-prone tasks.[1] Although single-investigator data extraction with secondary verification requires, on average, 107 minutes per study [2], methodological evaluations identified extraction errors in up to 67% of meta-analyses.[3] Common errors include data omissions, statistical misclassifications, misinterpretations of ambiguous primary studies, and data entry mistakes.

The use of artificial intelligence (AI) can potentially increase efficiency and correctness of the data extraction process.[4] Particularly, the introduction of large language models (LLMs), such as Generative Pre-trained Transformer (GPT)[5] or Claude,[6] has opened new possibilities for semi-automated data extraction, which combines machine-learning capabilities with human oversight. Unlike earlier natural language processing technologies used for data extraction, pre-trained LLMs can perform tasks without requiring task-specific training data, making them more accessible to users without technical expertise.

Initial evaluations of LLMs for data extraction demonstrated variable accuracy, ranging from 72% to 100%, compared with human reference standards.[7–12] However, their reliance on controlled experimental conditions and the use of pre-existing review datasets as benchmarks limit their generalizability to real-world applications. Furthermore, these studies assessed fully automated approaches without human involvement and evaluated the LLMs outside of the actual workflow of an evidence synthesis. While LLMs can potentially save time in data extraction, defining prompts (i.e., the specific inputs provided to a generative AI system to elicit a response) and interacting with the models add additional tasks to the review process. Therefore, validating their use within real-world workflows, specifically measuring both accuracy and time spent, is essential.

Our study aimed to validate a prospective workflow for an AI-assisted, semi-automated data extraction approach in which an LLM replaced a human investigator during the initial data extraction. We selected a semi-automated approach, as a fully automated method without human oversight is unlikely to be adopted for evidence synthesis in the near future.

## Methods

We registered the protocol of this study under: SWAR28 Gerald Gartlehner (2023 FEB 11 2102).pdf (qub.ac.uk).

### Research Questions

Our study was guided by two research questions:

1. What is the concordance between human-only and AI-assisted, semi-automated data extraction processes when used in real-world systematic review workflows, and how does the time-on-task compare between these two methods?
2. What is the accuracy of the AI-assisted data extraction process, and how do the types of errors compare to those of the human-only data extraction?

### Study Design

This study utilized a Study Within a Review (SWAR) [13] design, incorporating six ongoing systematic reviews [14–19] conducted by the Agency for Healthcare Research and Quality’s Evidence-based Practice Center Program. The systematic reviews addressed diverse topics and included both randomized and non-randomized studies. Except for one review, each systematic review was enrolled in the SWAR after its review team had completed the literature screening phase. If a review included more than 20 studies, an investigator not involved in the review randomly selected 20 studies for the SWAR. Due to delays, one review included only seven randomly selected studies.

For each participating systematic review, we conducted a prospective, parallel-group comparison of two data extraction processes, as outlined in Figure 1. Investigators of each review organized two independent data extraction teams. Team 1 (human-only data extraction) consisted of investigators who performed the initial data extraction from the original study report and other investigators who reviewed and revised the extracted information for completeness and correctness against the original study report. Team 2 (AI-assisted data extraction) uploaded the full-text Portable Document Format (PDF) of each study to the LLM Claude. They employed Claude for initial data extraction, followed by a human investigator who reviewed and revised the information for completeness and correctness against the study report. The teams consisted of different individuals, all with prior but varying experience in data extraction and a thorough understanding of the review’s topic and objectives.

**Figure 1:**
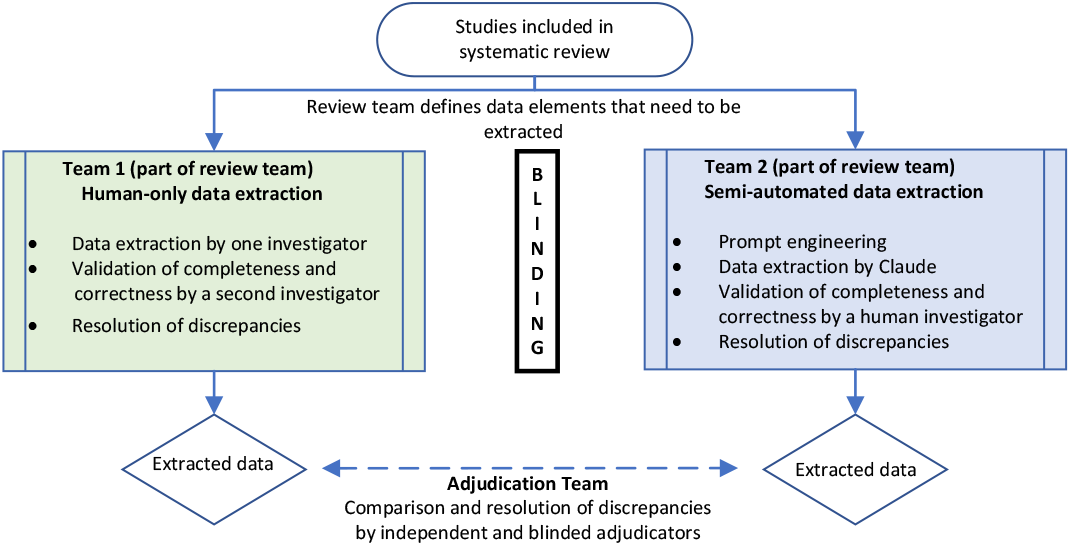
Outline of study design comparing two data extraction processes.

To ensure consistency and refine the process, both teams participated in an initial pilot data extraction using the same two studies. The pilot aimed to align team procedures, enhance comprehension of the data extraction tasks, and test the data extraction forms. Team 2 used this phase to develop and refine prompts for the LLM. After the pilot phase, the teams independently extracted data from the same selected studies, without having access to the other team’s extracted data.

Blinded adjudicators, uninvolved in the data extraction process or any of the six review teams, compared the results from the two data extraction teams (referred to as the adjudication team in Figure 1). In cases of discrepancies between extraction teams, two adjudicators independently reviewed the original study reports to determine which extracted data were accurate. A third, senior adjudicator, resolved any discrepancies and inconsistencies between the initial adjudicators.

### Prompt Engineering and Output Generation

We selected ClaudePro (versions 2.1, 3.0 Opus, and 3.5 Sonnet) for data extraction. According to Anthropic’s official policies, ClaudePro does not use uploaded PDFs or user interactions to train its models unless explicitly permitted, which respects the intellectual property rights of copyrighted study reports. Investigators received a training session on the use of Claude and the basics of prompt engineering. Prompt engineering involves designing text inputs (prompts) with the objective of accurate and succinct output from LLMs. Investigators received a pool of example prompts to work with, which were successfully used during a proof-of-concept study for data extraction with Claude 2.0[7]. They were informed that iteration on the prompt text is necessary for obtaining accurate results. Specifically, during the pilot phase, initial prompts were generated based on typical guidance that would be given to human extractors, and the resulting output was assessed. Based on the correctness and completeness of the output, adjustments were made to refine the prompts further. Throughout the study, investigators had access to a data scientist with expertise in prompt engineering when support was needed. Supplement 1 presents prompts used for each review.

### Outcomes and Error Analysis

The primary outcomes of the study were the concordance of data extracted by the two approaches and the time required to complete the data extraction for each study report. Concordance was defined as ‘*the proportion of data items that are factually congruent between the two methods, regardless of differences in style or presentation*’. The time required for data extraction included the total time spent on all tasks necessary for completion and verification, including prompt engineering in the AI-assisted process.

The secondary outcomes were the accuracy, recall, precision, and F1 score of the AI-assisted approach (see textbox for definitions) and the types of errors made by each of the data extraction approaches. To calculate the accuracy metrics of the AI-assisted approach, we used the finalized data by the adjudication team as the reference standard as it can be considered an enhanced version of human data extraction.

Two adjudicators independently classified error types (e.g., missed data, misallocated data, fabricated data) and the potential impact of errors (inconsequential, minor, or major). A third adjudicator resolved any discrepancies in adjudicator classifications.

#### Textbox

Definitions of performance metrics

**False negatives (FN):** The number of data items missed or incorrectly extracted from the study report.

**False positives (FP):** The number of data items for which data were extracted when no data were available in the study report (i.e., fabricated or hallucinated data).

**True negatives: (TN)** The number of data items that were correctly identified as not available in the study report.

**True positives (TP):** The number of data items correctly extracted from the study report.

**Accuracy:** The proportion of data items that were either correctly extracted from the study report or correctly noted as missing:

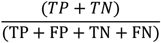

**Precision (=positive predictive value):** Of all extracted data items, the proportion that were correctly extracted:

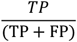

**Recall (=sensitivity):** Of all data items reported in a study report, the proportion that were correctly extracted:

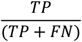

**F1-score:** An evaluation metric that combines the precision and recall (via harmonic mean) into a single proportion that ranges between 0 (poor) and 1 (perfect):

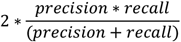

### Sample Size Calculation and Data Analysis

Our proof-of-concept study found an 83% concordance between human data extraction and LLM-conducted data extraction.[7] For the current study, we aimed to estimate the proportion of concordant data item pairs across four categories: study characteristics, participant characteristics, interventions and participant flow, and outcomes. To achieve a two-sided 95% Clopper-Pearson confidence interval that is approximately 7 percentage points wide, we required 500 data items for an observed binomial proportion of 83%.

To assess the time required for data extraction, investigators tracked their time while working on any of the tasks of the two data extraction processes. For team 2, this included the development and refinement of the prompts.

The unit of the analyses was the extracted data item as defined in each systematic review. We excluded data items that were subordinate to missed primary data items. For instance, if an entire outcome was missed by one of the extraction methods, the missed outcome was recorded as an error. Subordinate elements of the missed outcome—such as the point estimate, confidence interval, p-value, or length of follow-up—were not counted as additional errors.

In line with sample size considerations, we present proportions along with 95% Clopper-Pearson confidence intervals for concordance and accuracy, as well as for precision and recall, where the number of true positives is considered binomial with the sample size equal to the corresponding denominator.

For time on task, and types and severities of errors, we conducted descriptive analyses. Because of the exploratory nature of this study, we made no adjustments for multiplicity in the analyses. This allowed us to explore the data comprehensively and generate valuable insights without imposing strict adjustments for multiple comparisons.

## Results

The six systematic reviews included in the SWAR contributed 9,341 data elements extracted from 63 studies. Three reviews focused solely on RCTs, while three others included randomized and non-randomized studies (Table 1).

**Table 1:**
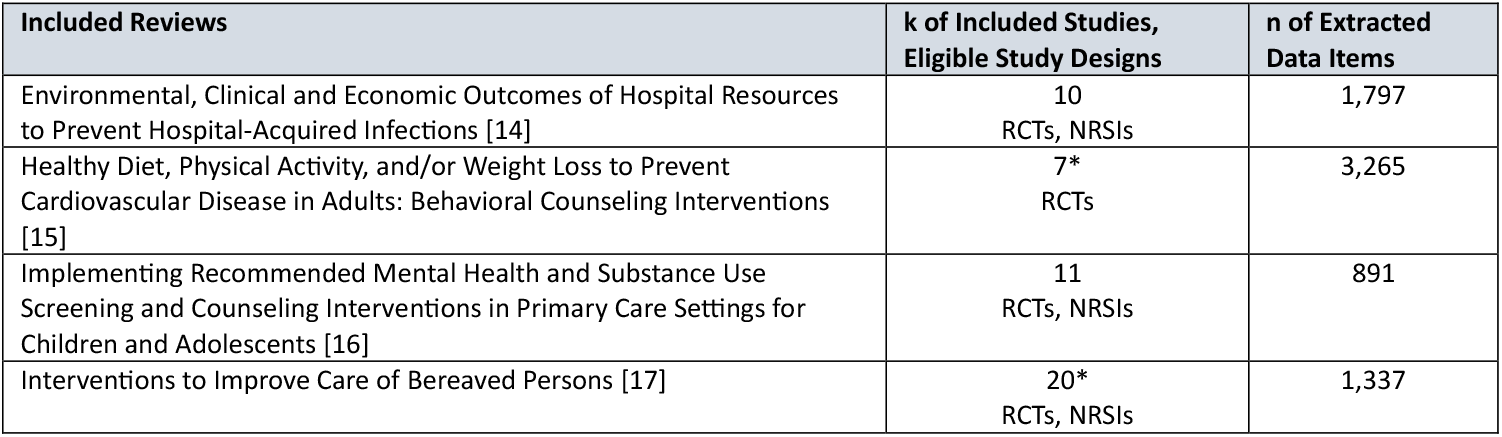
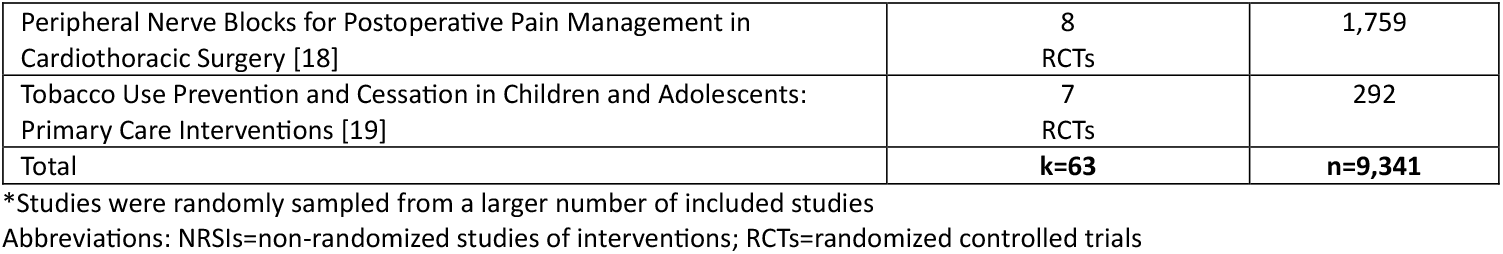
Topics, eligible study designs, number of contributing studies and data items of the contributing systematic reviews.

| Included Reviews | k of Included Studies, Eligible Study Designs | n of Extracted Data Items |
| --- | --- | --- |
| Environmental, Clinical and Economic Outcomes of Hospital Resources to Prevent Hospital-Acquired Infections [14] | 10<br>RCTs, NRSIs | 1,797 |
| Healthy Diet, Physical Activity, and/or Weight Loss to Prevent Cardiovascular Disease in Adults: Behavioral Counseling Interventions [15] | 7*<br>RCTs | 3,265 |
| Implementing Recommended Mental Health and Substance Use Screening and Counseling Interventions in Primary Care Settings for Children and Adolescents [16] | 11<br>RCTs, NRSIs | 891 |
| Interventions to Improve Care of Bereaved Persons [17] | 20*<br>RCTs, NRSIs | 1,337 |
| Peripheral Nerve Blocks for Postoperative Pain Management in Cardiothoracic Surgery [18] | 8<br>RCTs | 1,759 |
| Tobacco Use Prevention and Cessation in Children and Adolescents: Primary Care Interventions [19] | 7<br>RCTs | 292 |
| Total | **k=63** | **n=9,341** |

\*Studies were randomly sampled from a larger number of included studies
Abbreviations: NRSIs=non-randomized studies of interventions; RCTs=randomized controlled trials

### Concordance between data extraction approaches

The overall concordance between the two data extraction approaches was 77.2% (95% CI 76.3% to 78.0%, Table 2). Concordance was highest for data describing study characteristics (83.3%; 95% CI 80.3% to 86.1%), and lowest for data on outcomes and results (75.9%; 95% CI 74.8% to 76.9%; Figure 2). Levels of concordance varied from 65.5% to 88.4% across the six individual reviews (3).

**Table 2:** Performance metrics of the AI-assisted approach for each review and the SWAR overall.

| Review | Concordance (%)<br>(95% CI) | Accuracy (%)<br>(95% CI) | Recall (%)<br>(95% CI) | Precision (%)<br>(95% CI) | F1 score |
| --- | --- | --- | --- | --- | --- |
| Environmental, Clinical and Economic Outcomes of Hospital Resources to Prevent Hospital-Acquired Infections [14] | 65.5<br>(63.2-67.7) | 88.2<br>(86.6-89.7) | 86.3<br>(84.4-87.9) | 99.8<br>(99.4-100) | 0.93 |
| Healthy Diet, Physical Activity, and/or Weight Loss to Prevent Cardiovascular Disease in Adults: Behavioral Counseling Interventions [15] | 85.1<br>(83.8-86.3) | 93.5<br>(92.6-94.3) | 94.3<br>(93.5-95.1) | 98.9<br>(98.6-99.1) | 0.96 |
| Implementing Recommended Mental Health and Substance Use Screening and Counseling Interventions in Primary Care Settings for Children and Adolescents [16] | 88.4<br>(86.2-90.4) | 94.5<br>(92.8-95.9) | 88.0<br>(84.5-91.0) | 100<br>(99.0-100) | 0.94 |
| Interventions to Improve Care of Bereaved Persons [17] | 68.6<br>(66.0-71.1) | 86.4<br>(84.4-88.2) | 86.1<br>(84.0-88.0) | 99.2<br>(98.4-99.6) | 0.92 |
| Peripheral Nerve Blocks for Postoperative Pain Management in Cardiothoracic Surgery [18] | 76.4<br>(74.4-78.4) | 92.2<br>(90.8-93.4) | 84.1<br>(81.2-86.7) | 96.3<br>(94.6-97.7) | 0.90 |
| Tobacco Use Prevention and Cessation in Children and Adolescents: Primary Care Interventions [19] | 69.9<br>(64.2-75.1) | 83.9<br>(79.2-87.9) | 81.0<br>(75.6-85.7) | 100<br>(98.2-100) | 0.89 |
| **Combined across reviews** | **77.2**<br>**(76.3-78.0)** | **91.0**<br>**(90.4-91.6)** | **89.4**<br>**(88.6-90.1)** | **98.9**<br>**(98.6-99.1)** | **0.94** |

Abbreviations: CI=confidence interval

**Figure 2:**
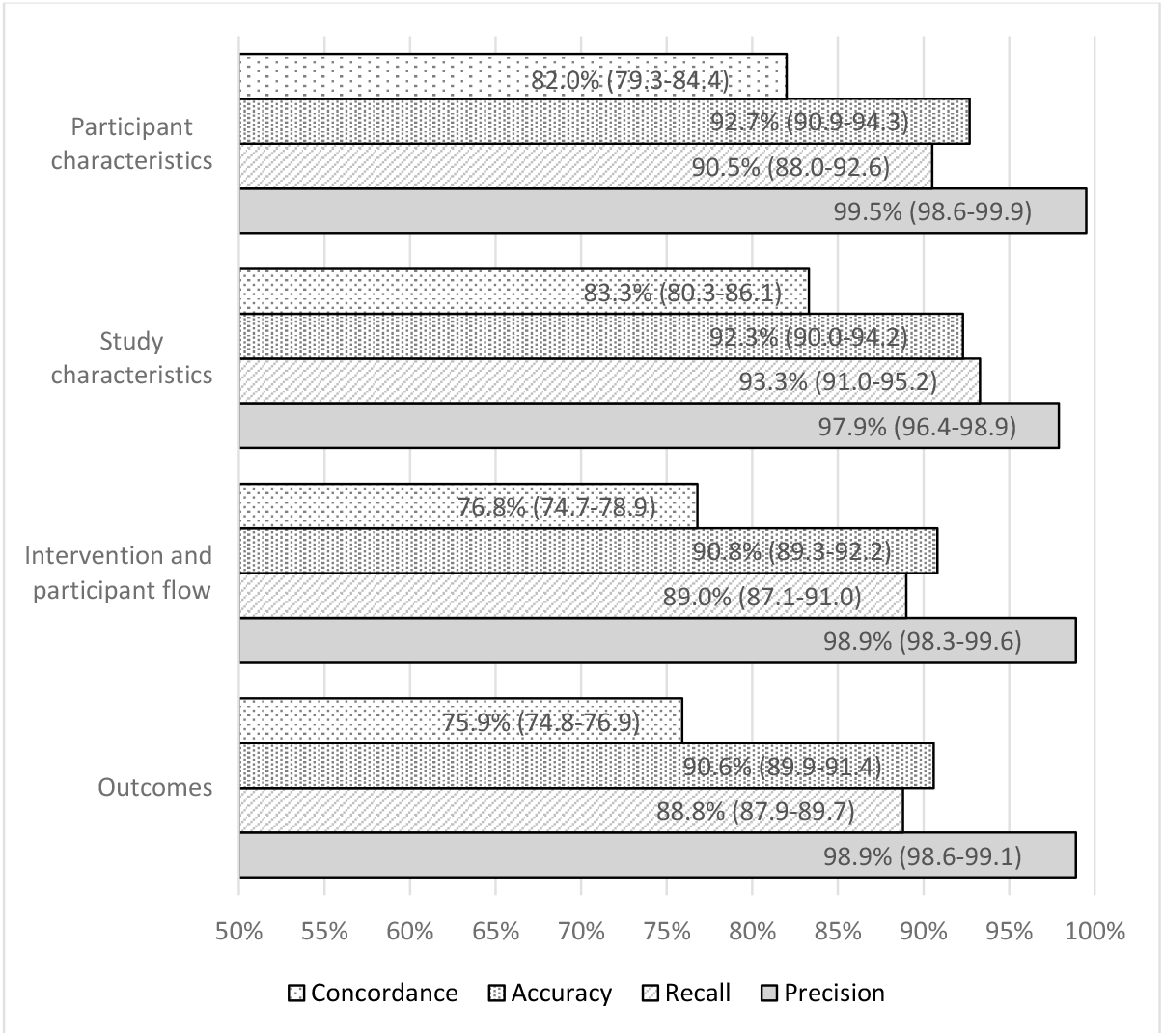
Concordance, accuracy, recall, and precision by data categories. illustrates concordance, accuracy, recall, and precision stratified by different data categories. Overall, performance metrics were consistent for different data categories, except for higher concordance observed for participant and study characteristics than for interventions and participant flow, or outcomes and results.

Of the 9,341 extracted data item pairs, 2,132 were discordant between human-only and AI-assisted extractions. When compared to the adjudicated reference standard, human-only extractions were incorrect in 41.7% (n = 890) of the discordant items, while AI-assisted extractions were incorrect in 32.9% (n = 701). In some instances, both approaches had incorrect extractions (6.6%; n = 140) when compared to the adjudicated reference standard. In 18.8% (n = 401) of discordant pairs, neither extraction was wrong. Such situations arose when the reason for discordance was the level of reported detail, when data elements were poorly defined, or prompts were ambiguous. For instance, one approach might have extracted intention-to-treat results while the other extracted per-protocol results, due to an unclear definition of the data item or a prompt that did not specify which results to extract.

### Performance of AI-assisted Data Extraction

The AI-assisted data extraction approach demonstrated an overall accuracy of 91.0% (95% CI 90.4% to 91.6%, Table 2) when compared to the adjudicated reference standard. Accuracy refers to the percentage of data items correctly identified, either as accurate extractions when data were available or reported as absences when data were missing. Recall, which measures the percentage of accurately extracted data items by the AI-assisted approach, out of all available data items in study reports, was 89.4% (95% CI 88.6% to 90.1%). Precision, indicating the percentage of correctly extracted data items, out of all extracted data items, was 98.9% (95% CI 98.6% to 99.1%). The F1 score, representing the harmonic mean of recall and precision, was 0.94. Table 2 also presents performance metrics for each included review.

### Error Analysis

Table 3 describes the different error types, the potential impact of errors, and the frequency of each error type out of all extracted items. Among the 9,341 extracted data items, errors were identified by the adjudicated reference standard in 1,731 cases (AI-assisted 9.0% [95% CI 8.4% to 9.6%] vs. human-only 11.0% [95% CI 10.4% to 11.7%]). The risk of major errors—those that significantly compromise data accuracy and could lead to erroneous conclusions if left uncorrected—was nearly identical between the two groups (AI-assisted 2.5% [2.2% to 2.8%] vs. human-only 2.7% [2.4% to 3.2%]). The most common errors for both approaches were missed data (AI-assisted 5.5% vs. human-only 6.2%) and misallocated data (AI-assisted 1.8% vs. human-only 1.9%). Fabricated data, which refers to data items that were not present in source study reports but that were captured by human data extractors or erroneously generated by the LLM (hallucinated) and not corrected by the human reviewer, were rare in both approaches (AI-assisted 0.8% vs. human-only 0.5%).

**Table 3:** Types and frequencies of errors and their potential impacts.

| Types of error | Definitions | Number of Errors and Percentage of all Extracted Data Items (n=9,341) |  |
| --- | --- | --- | --- |
|  |  | Human-only approach* | AI-assisted approach* |
| Missed or omitted data | Data available in the source study report that were either missed or omitted by the data extraction process. | 578 (6.2%) | 516 (5.5%) |
| Misallocated data | Data provided in the source study report that were allocated to the wrong data extraction field. | 181 (1.9%) | 170 (1.8%) |
| Fabricated data | Data not available in the source study report that were captured by human data extractors or erroneously generated (hallucinated) by the LLM and not corrected by the human reviewer. | 48 (0.5%) | 72 (0.8%) |
| Difference in level of detail** | The level of detail captured is the only difference between data extractions. | 47 (0.5%) | 37 (0.4%) |
| Incorrect calculations | Mathematically incorrect calculations of data elements based on information provided in the source study report, including rounding errors. | 20 (0.2%) | 15 (0.2%) |
| Other | Optional “other” field if none of the above apply, for example typos or obvious misunderstandings of instructions | 156 (1.7%) | 31 (0.3%) |
| **Potential impact of error** |  |  |  |
| Major error | This error significantly compromises the correctness of the data, and, if uncorrected, could lead to erroneous conclusions; for example, grossly incorrect calculations, misallocated data that results in a different interpretation, or fabricated data that result in a new or different interpretation of the data. | 255 (2.7%) | 231 (2.5%) |
| Minor error | This error is less severe than a major error and may or may not impact interpretation of the existing data; for example, small calculation errors or rounding errors that do not critically affect the data's overall utility, additional or alternative language describing study inclusion/exclusion criteria or the intervention that don't alter the meaning in a significant way. | 683 (7.3%) | 496 (5.3%) |
| Inconsequential difference | This difference most likely would not impact the interpretation of the data, for example differences in the level of detail of inclusion criteria. | 92 (1.0%) | 114 (1.2%) |

\*When both approaches produced incorrect extractions for the same data item, errors were attributed to each approach independently. As a result, the total number of recorded errors exceeds the total number of incorrectly extracted data items.
\*\* Difference in level of detail is not an error per se but a discrepancy that influenced concordance.

### Time on Task

Across the six included reviews, the AI-assisted approach reduced the median time required for data extraction by 41 minutes per study compared to human-only extraction (84 vs. 125 minutes). The range of time differences across the six reviews spanned from 78 minutes shorter to 25 minutes longer (Figure 3). In five of the six reviews, data extraction with AI assistance required less time than human-only efforts.

**Figure 3:**
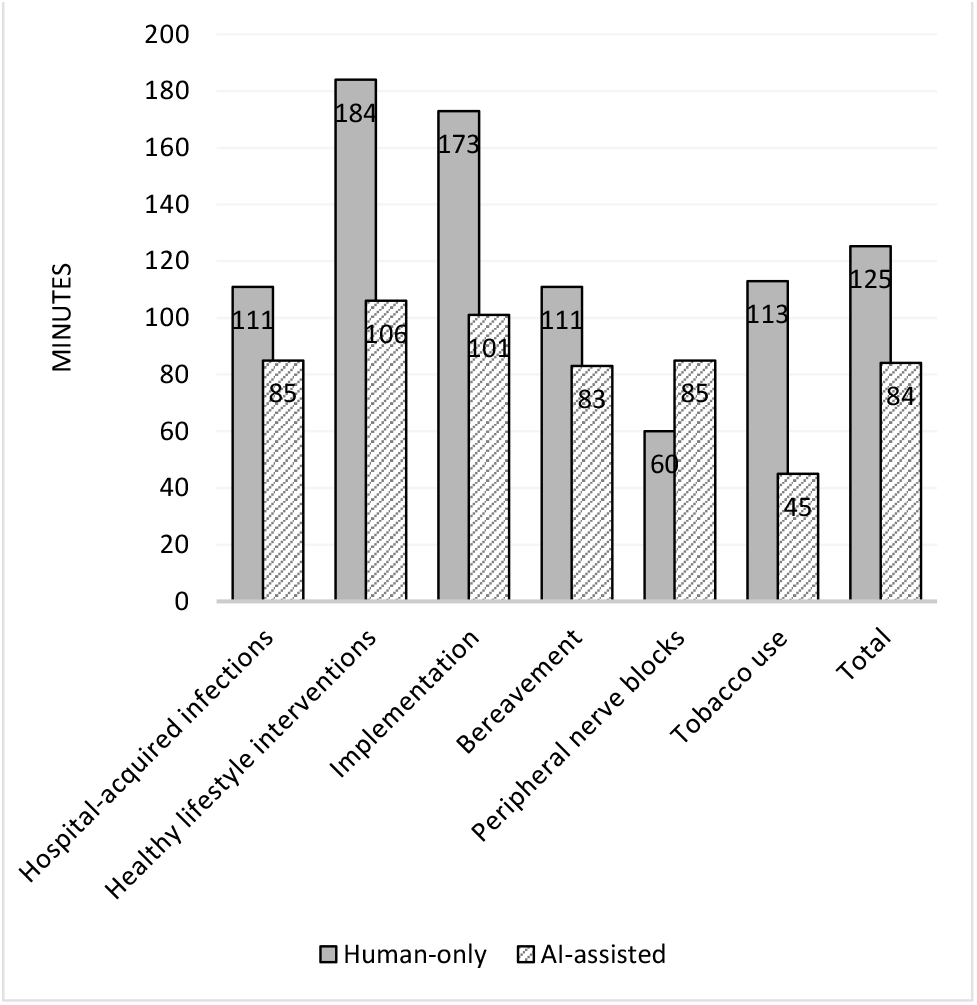
Median time spent for data extraction per study

### Additional Exploratory Analyses

We investigated whether the basic data types of the extracted information influenced concordance and performance metrics. We categorized the data into three types: numerical, string, and mixed data. For the purpose of this study, numerical data referred to data in which numbers provided the primary information for extraction, even if characters were also included (e.g., 100 participants). String data, on the other hand, involved information where characters, words, or sentences were most relevant, regardless of the presence of numerals (e.g., follow-up with 12-lead electrocardiograph). Mixed data consisted of both numerals and characters, where information from both were equally important (e.g., drug dosage 20 mg). Overall, extractions of string data resulted in higher concordance and better accuracy and recall compared with numerical or mixed data types (<u>Appendix</u> Figure 1).

## Discussion

Our SWAR utilized six ongoing systematic reviews and is the first study to prospectively evaluate the workflow of AI-assisted data extraction under real-world settings. Compared with the traditional human-only extraction method, the AI-assisted approach yielded encouraging results, with fewer incorrect extractions (9.0% vs. 11.0%), comparable risks of major data extraction errors (2.5% vs. 2.7%), and a median time saving of 41 minutes per study. The concordance between the two extraction processes was 77.2% overall, highest for study characteristics data and lowest for outcomes data.

Our study also confirmed previous work [3; 20] that the traditional human-only data extraction is error-prone. Although all extractions were performed by professional reviewers and checked for correctness and completeness by a second investigator, 11% of the data items extracted by humans contained one or more errors. Most of these errors were minor and inconsequential, yet they highlight the fallible nature of human data extraction. This was further confirmed by the AI-assisted teams’ performance. While the AI-assisted teams had, on average, 9% incorrect extractions—fewer than the human-only teams—most errors in the AI-assisted teams were missed data items. They were overlooked by both the LLM during the initial extraction and the human investigator who reviewed the LLM’s output.

Because our study is the first workflow validation of AI-assisted data extraction, no direct comparison of findings with other studies is possible. Existing model validations of LLMs for data extraction reported accuracies ranging from 72% to 100%, consistent with the accuracy metrics reported in our SWAR.[7–10; 12]

Our study has several notable strengths. A primary strength is its design as a workflow validation study with a prospective design. Compared with model validation studies that examine the performance and reliability of an LLM under specific, controlled conditions using already existing datasets, workflow validation studies integrate the use of the LLM into the workflow of an ongoing review, providing a detailed perspective on the model’s practical effectiveness, efficiency, and utility.[21] The prospective design protects against data contamination, which can arise when the dataset used to evaluate performance also contributed to LLM training, potentially inflating its capabilities.[22] Given that developers of LLMs rarely disclose their training datasets, the prospective design of our study utilizing ongoing, unpublished reviews provides a rigorous safeguard against this risk.[21]

Additionally, to protect against benchmark bias, a form of classification bias which can arise when the reference standard is imperfect, we did not consider the human-only extraction as the reference standard. Instead, we verified each discrepancy between human-only and AI-assisted extractions against the original study report by blinded adjudicators. This allowed for a more accurate assessment of the performance of both approaches.

Another notable strength is the inclusion of a wide range of systematic review topics and study designs, encompassing both RCTs and NRSIs. This diversity enhances the study’s generalizability across different research contexts, especially given that performance metrics showed no substantial variation across the individual reviews. Finally, the large dataset, comprising 9,341 data item pairs, ensures sufficient statistical power for robust analyses.

Despite the methodological strengths, our study also has several limitations. As with any workflow validation studies, the generalizability of the findings to other types of data and evidence synthesis may be restricted. While the six included reviews covered a broad range of topics, all focused on healthcare interventions. Results could differ for reviews of diagnostic test accuracy, prognostic studies, or studies in other sectors (e.g., education, economics, business). Another limitation stems from human variation. Differences in systematic review experience, levels of detail in data extraction, team approaches to validating data extractions, and proficiency in engineering prompts may influence the reproducibility of our results. These factors inherently limit the broader applicability of the findings.

Although time spent on task was one of our primary outcomes, consistently tracking time across reviews was challenging. For example, rate restrictions of the LLM due to factors like institutional traffic varied across institutions and limited the workflow at one institution. Ultimately, we assumed that time tracking within each review was conducted under similar conditions, and that the absolute differences in time spent on data extraction reflected actual results within each review. However, the comparability of time spent across reviews may be limited. Nonetheless, the consistent time savings observed in the AI-assisted approach in five out of six reviews strengthen confidence in the overall direction of the results.

Assessing the concordance of data extractions and classifying errors often required subjective judgment. To limit subjectivity, two study investigators from the adjudication team independently verified errors, classified their type and severity, and a third investigator resolved any discrepancies between them. Nevertheless, inconsistencies in classifications are likely, and different investigators may have reached different conclusions regarding the types and severities of errors. Although we attempted to blind the adjudication team to the data extraction approach, in some cases it was apparent which extraction was from the AI-assisted approach due to the verbose nature of the responses, which sometimes included irrelevant information. Furthermore, since we adjudicated only discordant data extractions, we have missed instances where extraction results were concordant but both extractions were incorrect. However, we assumed that such cases are rare.

Due to the rapid advancement of LLMs, we switched from Claude 2 to Claude 3 during the SWAR. This change was in line with our protocol but may have contributed to differences in performance metrics across reviews.

Finally, although our adjudicators assessed the *potential* impact of errors, we did not assess the *actual* impact of data extraction errors on systematic review results and conclusions. The consequences of inaccuracies in different data elements can vary, potentially impacting a review’s conclusions differently. In our study, the adjudicators who judged the seriousness of the error were experienced systematic reviewers but were not part of the teams for each systematic review.

Although the results of our study are encouraging, several key areas warrant further investigation to advance the field. Our research focused on a single scenario where the LLM performed the initial data extraction, and human investigators verified its work. However, many alternative scenarios could be explored. For instance, overall performance might improve if the roles were reversed, with an LLM verifying human data extraction. Alternatively, a second LLM could review the work of the first LLM before human verification. Ultimately, a prospective workflow validation of extraction by LLM alone without human review would help to identify where humans must stay involved. Any advancement in the application of LLMs to the data extraction process also needs to be validated. In this study, investigators manually developed and entered prompts into the LLM’s web-based user interface. Developers of systematic review software are already working to integrate LLMs directly into software applications. Such integration could provide greater time savings and improved performance but might reduce flexibility if users are unable to adapt prompts to their specific review needs. Developing standardized prompts for common data items could enhance LLM performance and ensure consistency across various contexts. In-depth case studies comparing LLM-assisted and human-only methodologies should extend beyond assessing the correctness of data extraction. They should also examine how inaccuracies impact evidence synthesis, such as meta-analyses, and influence the ultimate conclusions of systematic reviews.

Future research also needs to develop methods to assess the extent to which data contamination can bias LLM evaluations. For example, developing a private benchmark dataset [23] (i.e., where each data item is annotated with specific labels and the dataset remains inaccessible to models during evaluation) based on full-text study reports from systematic reviews, could provide a standardized reference for accuracy assessments. Such a dataset would also eliminate reliance on human data extraction as a reference standard.

## Data Availability

Statistical code and data set: Available from Claus Nowak.

## Disclaimer

This project was funded under Contract No. 75Q80120D00007 Task Order 75Q80120F32002 from the Agency for Healthcare Research and Quality (AHRQ), U.S. Department of Health and Human Services (HHS). The authors of this manuscript are responsible for its content. Statements in the manuscript do not necessarily represent the official views of or imply endorsement by AHRQ or HHS.

## Acknowledgments

The authors thank Ian Thomas and Amanda Konet from RTI International, as well as Elizabeth Webber from the Kaiser Permanente Center for Health Research, for their support with prompt engineering. They also extend their gratitude to Claus Nowak from the University for Continuing Education Krems, Austria, for the statistical analyses. We are grateful to the following individuals for their contributions to data extraction for the individual systematic reviews included in this study: Sheila Patel and Els Houtsmuller from RTI International; Shelley Selph, Miranda Pappas, Rebecca Holmes, Steffani Bailey, and Erica Hart from the Oregon Health and Science University; Benjamin Rouse from ECRI; Mary Butler and Toyin Lamina from the University of Minnesota; and Carrie Patnode from the Kaiser Permanente Center for Health Research.

We would like to acknowledge the US Agency for Healthcare Research and Quality EPC Program, the RTI International Innovation Fund, and the RTI International Fellow’s Program for support of this project.

## Reproducible Research Statement

Statistical code and data set: Available from Claus Nowak.

## Appendix

**Appendix Figure 1:**
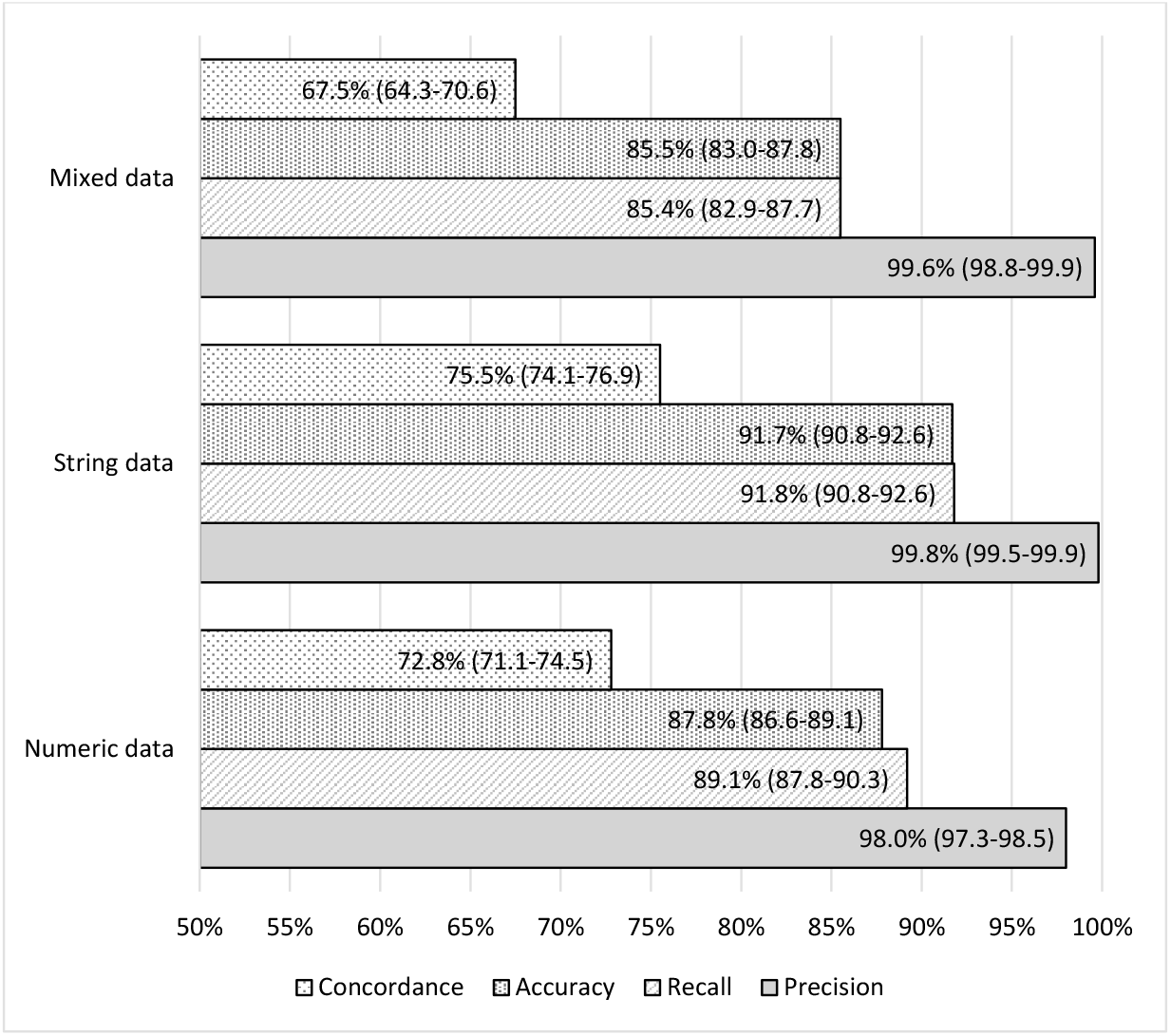
Concordance, accuracy, recall, and precision stratified by basic data type

## References

1. Nussbaumer-Streit B, Ellen M, Klerings I, Sfetcu R, Riva N, Mahmić-Kaknjo M, et al. Resource use during systematic review production varies widely: a scoping review. Journal of clinical epidemiology. 2021;139:287–96.

2. Li T, Saldanha IJ, Jap J, Smith BT, Canner J, Hutfless SM, et al. A randomized trial provided new evidence on the accuracy and efficiency of traditional vs. electronically annotated abstraction approaches in systematic reviews. Journal of Clinical Epidemiology. 2019;115:77–89.

3. Mathes T, Klassen P, Pieper D. Frequency of data extraction errors and methods to increase data extraction quality: a methodological review. BMC Med Res Methodol. 2017;17(1):152.

4. Schmidt L, Finnerty Mutlu AN, Elmore R, Olorisade BK, Thomas J, Higgins JPT. Data extraction methods for systematic review (semi)automation: Update of a living systematic review. F1000Res. 2021;10:401.

5. OpenAI R. GPT-4 technical report. arXiv, 2303-08774. 2023.

6. Anthropic. Claude: https://www.anthropic.com/claude (Accessed: December 14, 2024).

7. Gartlehner G, Kahwati L, Hilscher R, Thomas I, Kugley S, Crotty K, et al. Data extraction for evidence synthesis using a large language model: A proof-of-concept study. Res Synth Methods. 2024;15(4):576–89.

8. Khraisha Q, Put S, Kappenberg J, Warraitch A, Hadfield K. Can large language models replace humans in systematic reviews? Evaluating GPT-4’s efficacy in screening and extracting data from peer-reviewed and grey literature in multiple languages. Research Synthesis Methods. 2024;15(4):616–26.

9. Konet A, Thomas I, Gartlehner G, Kahwati L, Hilscher R, Kugley S, et al. Performance of two large language models for data extraction in evidence synthesis. Research Synthesis Methods. 2024;15(5):818–24.

10. Mahmoudi H, Chang D, Lee H, Ghaffarzadegan N, Jalali MS. A Critical Assessment of Large Language Models for Systematic Reviews: Utilizing ChatGPT for Complex Data Extraction. Available at SSRN 4797024. 2024.

11. Panayi A, Ward K, Benhadji-Schaff A, Ibanez-Lopez AS, Xia A, Barzilay R. Evaluation of a prototype machine learning tool to semi-automate data extraction for systematic literature reviews. Systematic Reviews. 2023;12(1):187.

12. Motzfeldt Jensen M, Brix Danielsen M, Riis J, Assifuah Kristjansen K, Andersen S, Okubo Y, et al. ChatGPT-4o can serve as the second rater for data extraction in systematic reviews. PLoS One. 2025;20(1):e0313401.

13. Devane D, Burke NN, Treweek S, Clarke M, Thomas J, Booth A, et al. Study within a review (SWAR). Journal of Evidence-Based Medicine. 2022;15(4):328–32.

14. Effective Health Care Program, Agency for Healthcare Research and Quality. Research Protocol: Environmental, Clinical and Economic Outcomes of Hospital Resources to Prevent Hospital-Acquired Infections: https://effectivehealthcare.ahrq.gov/products/prevent-hai/protocol updated August 2024 (Accessed: December 14, 2024).

15. U.S. Preventive Services Task Fore. Final Research Plan: Healthy Diet, Physical Activity, and/or Weight Loss to Prevent Cardiovascular Disease in Adults: Behavioral Counseling Interventions: https://www.uspreventiveservicestaskforce.org/uspstf/document/final-research-plan/behavioral-counseling-interventions-promote-healthy-diet-physical-activity-weight-loss-prevent-cardiovascular-disease-adults updated November 02, 2023 (Accessed: December 14, 2024).

16. Agency for Healthcare Research and Quality. Research Protocol: Implementation of Recommended Screening and Counseling Interventions to Prevent Mental Health Disorders in Children and Adolescents: https://effectivehealthcare.ahrq.gov/products/behavioral-health-screening/protocol updated December 08, 2023 (Accessed: December 14, 2024).

17. Agency for Healthcare Research and Quality. Research Protocol: Interventions to Improve Care of Bereaved Persons: https://effectivehealthcare.ahrq.gov/products/bereaved-persons/protocol updated December 04, 2023 (Accessed: December 14, 2024).

18. Agency for Healthcare Research and Quality. Research Protocol: Peripheral Nerve Blocks for Postoperative Pain Management in Cardiothoracic Surgery: https://effectivehealthcare.ahrq.gov/products/peripheral-nerve-blocks/protocol updated May 17, 2024 (Accessed: December 14, 2024).

19. U.S. Preventive Services Task Fore. Final Research Plan: Tobacco Use Prevention and Cessation in Children and Adolescents: Primary Care Interventions: https://www.uspreventiveservicestaskforce.org/uspstf/document/final-research-plan/tobacco-use-intervention-prevention-cessation-children-adolescents updated June 27, 2024 (Accessed: December 14, 2024).

20. Xu C, Yu T, Furuya-Kanamori L, Lin L, Zorzela L, Zhou X, et al. Validity of data extraction in evidence synthesis practice of adverse events: reproducibility study. Bmj. 2022;377:e069155.

21. Gartlehner G, Kahwati L, Nussbaumer-Streit B, Crotty K, Hilscher R, Kugley S, et al. From promise to practice: challenges and pitfalls in the evaluation of large language models for data extraction in evidence synthesis. BMJ Evidence-Based Medicine. 2024:bmjebm-2024-113199.

22. Carlini N, Ippolito D, Jagielski M, Lee K, Tramer F, Zhang C. Quantifying memorization across neural language models. arXiv preprint arXiv:220207646. 2022.

23. Rajore T, Chandran N, Sitaram S, Gupta D, Sharma R, Mittal K, et al. Truce: Private benchmarking to prevent contamination and improve comparative evaluation of llms. arXiv e-prints. 2024:2403.00393.

